# Ethnic disparities in hospitalisation and hospital-outcomes during the second wave of COVID-19 infection in east London

**DOI:** 10.1101/2021.07.05.21260026

**Authors:** Y I Wan, V J Apea, R Dhairyawan, Z A Puthucheary, R M Pearse, C M Orkin, J R Prowle

## Abstract

**Objectives:** To determine if changes in public behaviours, developments in COVID-19 treatments, improved patient care, and directed policy initiatives have altered outcomes for minority ethnic groups in the second pandemic wave.

**Design:** Prospectively defined observational study using registry data.

**Setting:** Four acute NHS Hospitals in east London.

**Participants:** Patients aged ≥16 years with an emergency hospital admission with SARS-CoV-2 infection between 1^st^ September 2020 and 17^th^ February 2021.

**Main outcome measures:** Primary outcome was 30-day mortality from time of index COVID-19 hospital admission. Secondary endpoints were 90-day mortality and need for ICU admission. Multivariable survival analysis was used to assess associations between ethnicity and mortality accounting for predefined risk factors. Age-standardised rates of hospital admission relative to the local population were compared between ethnic groups.

**Results:** Of 5533 patients, the ethnic distribution was White (n=1805, 32.6%), Asian/Asian British (n=1983, 35.8%), Black/Black British (n=634, 11.4%), Mixed/Other (n=433, 7.8%), and unknown (n=678, 12.2%). Excluding 678 patients with missing data, 4855 were included in multivariable analysis. Relative to the White population, Asian and Black populations experienced 4.1 times (3.77-4.39) and 2.1 times (1.88-2.33) higher rates of age-standardised hospital admission. After adjustment for various patient risk factors including age, sex, and socioeconomic deprivation, Asian patients were at significantly higher risk of death within 30 days (HR 1.47 [1.24-1.73]). No association with increased risk of death in hospitalised patients was observed for Black or Mixed/Other ethnicity.

**Conclusions:** Asian and Black ethnic groups continue to experience poor outcomes following COVID-19. Despite higher-than-expected rates of admission, Black and Asian patients experienced similar or greater risk of death in hospital, implying a higher overall risk of COVID-19 associated death in these communities.

**Strengths and limitations of this study:**

- This study represents one of the largest descriptors of outcomes in minority ethnic patients with COVID-19 distinguished by the majority ethnically diverse cohort within a catchment area of approximately one million people in the east of London.
- Large absolute numbers of patients drawn from a single geographic region and treated within the same hospital system minimize many of the geographic biases present within other studies including the impact of variation in transmission risk.
- Our analyses are strengthened by adherence to a prespecified analysis plan, inclusion of a detailed baseline, comorbidity, and COVID-19 risk factors in multivariable modelling, and sensitivity tests using different measures of comorbidity.
- However as with all observational studies, not all potential contributing risk factors could be assessed, including other measures of baseline health status such as nutritional or lifestyle influences as well as contextual factors such as household composition and occupation.
- We were not able to include suspected but not proven COVID-19 cases or community cases not requiring hospital admission or the effect of the differing viral strains in the first and second wave.

## INTRODUCTION

Multiple studies have described increased mortality in Black and Asian people with COVID-19 in the UK.^1-5^ In many of these reports, Black people were at equal or greater risk of death than Asian people. These studies have included large analyses of hospitalised patients and analyses of COVID-19 associated deaths within large sets of primary care records.^5^ However due to geographic variation, representation of ethnic minority groups in many of these datasets were small and this has potential to bias analyses. Our initial report of almost 2000 COVID-19 associated emergency admissions in the first wave of COVID-19 was distinguished by inclusion of a significant number of patients drawn from the same geographic region (east London) where more than fifty percent of patients came from ethnically diverse backgrounds.^6^

In the winter of 2020, the UK experienced a devastating second wave of COVID-19 which compared to the first, had greater impact on healthcare services and higher number of deaths.^7,8^ Since the first wave, there have been societal behavioural change, improved COVID-19 treatments, and better algorithms of care, which have the potential to both mitigate or magnify ethnic inequalities in adverse outcomes associated with COVID-19. Furthermore, there have been directed initiatives aimed at identifying the driving factors behind these disparities.^9^ However, it is unclear if these ethnic inequalities persisted over time. In the UK, east London was at the epicentre of both waves of COVID-19. In the second wave, across four acute NHS hospitals in this region, we continued to treat high acuity patients and centralised surge critical care capacity on one site in a purpose-built ICU (The Queen Elizabeth Unit). This large, regional dataset afforded extensive analyses of COVID-19 patients to further characterise the risk factors within different ethnic groups of hospitalised COVID-19 patients. In this study, we aimed to determine if ethnic disparities during the first wave of COVID-19 have been mitigated during the second wave and the long-term survival of the first wave patients after one year.

## METHODS

We considered all adults (age ≥16 years) with confirmed SARS-CoV-2 infection admitted as emergencies to one of the four acute hospitals within Barts Health NHS Trust between 1^st^ September 2020 and 17^th^ February 2021. The first emergency admission encompassing the first positive SARS-CoV-2 test on real-time polymerase chain reaction (PCR) test, or the first emergency admission within two weeks of positive outpatient testing was defined as the index admission. Community diagnoses without an associated emergency hospital admission were excluded. Patients with unknown or undisclosed ethnicity status were collected for comparison but excluded from primary statistical analyses. In addition, we conducted extended follow-up of the 1996 SARS-CoV-2 hospital admissions up to 20^th^ May 2020, presented previously, to assess one-year survival.^6^ Finally, for patients resident in the London Boroughs of Tower Hamlets, Waltham Forest, and Newham, we assessed the relative frequency of SARS-CoV-2 associated acute hospital admissions, compared to the age and ethnicity distribution of this local population in the 2011 UK census.

### Data sources

Clinical and demographic data, blood results, and coding data from current and prior clinical encounters, were collated from the Barts Health Cerner Millennium Electronic Medical Record (EMR) data warehouse by members of the direct clinical care team. Mortality data was available to 25^th^ May 2021, enabling 90-day follow-up. Population level analyses of age, sex, and ethnicity distributions in the London boroughs of Tower Hamlets, Newham,, and Waltham Forest were taken from the 2011 census provided by Official Labour Market Statistics (nomis).^10^

### Definition of key variables

Ethnicity was defined using the NHS ethnic category codes and based on five high-level groups: White, Asian or Asian British, Black or Black British, Mixed, and Other. In the NHS categorisation, the Asian group is predominantly South Asian (Indian, Bangladeshi, Pakistani) with Chinese ethnicity assigned to the Other group, while in the 2011 census Chinese is assigned to the Asian higher-level group, for consistency the NHS high-level group definitions were applied to the census data. Due to small numbers in the Mixed group, the Mixed and Other categories were merged in multivariable modelling to preserve statistical power. Relative measures of socioeconomic deprivation were assessed using the English Indices of Deprivation 2020 by matching patient postcode to the national Index of Multiple Deprivation (IMD) using the Office of National Statistics Postcode Directory.^11^ Due to the level of deprivation of the majority of our study population, relative deprivation in this study was analysed based on quintiles of IMD within our study population. Baseline comorbid diseases, Charlson Comorbidity Index (CCI), and Hospital Frailty Risk Score (HFRS) were identified by mapping to ICD-10 coding.^12^ Body Mass Index (BMI) was calculated by height and weight measurements taken at or during the immediately preceding admission episode. Rockwood Clinical Frailty Scoring (RFS) was assessed by the admitting medical team.^13^ Full definitions are detailed in supplementary materials.

### Outcomes

Primary outcome was 30-day mortality from time of index COVID-19 hospital admission. Secondary endpoints were 90-day mortality and need for ICU admission defined by need for advanced respiratory support alone or basic respiratory support together with support for at least two organ systems.

### Statistical analyses

Our analysis plan followed our first wave COVID study.^14^ Baseline characteristics are presented as mean and SD, median and IQR, or number and percentage, as appropriate. We compared proportions using Pearson’s χ^2^ test or Fisher’s exact test and continuous variables using Wilcoxon rank-sum test and Kruskal-Wallis rank sum test, as appropriate. Time-to-event analysis was undertaken with survivors censored at 30 or 90 days or at time of maximal follow-up. Prolonged follow-up of our first wave cohort was examined over 270 days. Cox-proportional hazards models was used to assess survival adjusted for age, sex, and predefined risk factors associated with adverse outcomes in COVID-19: IMD quintile, smoking status, obesity, diabetes, hypertension, and chronic kidney disease (CKD). Additional multivariable models were also carried out as sensitivity analyses using aggregate CCI as a measure of total comorbid disease burden, and HFRS at hospital admission. The proportional hazard assumption was assessed by inspection of scaled Schoenfeld residual plots and investigated by stratification if required.^15^ Factors associated with ICU admission were examined using logistic regression. Effect measures are presented as Hazard or Odds ratios with 95% CI. All analyses were performed using R v4.0.2 (R Core Team 2020).

### Rates of admission

Barts Health hospitals primarily serve three London Boroughs: Tower Hamlets, Newham, and Waltham Forest. Patients residing within these areas were identified by postcode and age-adjusted rates of admission per 100,000 of the local population within each ethnic group were derived for COVID-19 first and second wave admissions by standardisation to the Revised European Standard Population.^16^ For comparison to non-COVID admissions, we also calculated age-standardised admissions per 100,000 for each ethnic group in all Barts Health emergency admissions residing in these three boroughs during the years 2013-18 inclusive. Overall rates are presented relative to that of the White ethnic group for comparison of relative admission rates between first and second waves and the pre-COVID baseline.

## RESULTS

A total of 5533 patients were identified (Fig S1). The majority of admissions occurred during December 2020 and January 2021 (Fig S2). Three-quarters of patients were classified as being in the four most deprived socioeconomic deciles in England (Table S1). Only around one third were White (n=1805, 32.6%). Ethnically diverse groups comprised around 70% of the cohort: Asian or Asian British (n=1983, 35.8%), Black or Black British (n=634, 11.4%), Mixed/Other (n=433, 7.8%) and unknown or undisclosed (n=678, 12.2%).

### Population characteristics

Baseline characteristics, interventions, and outcomes across ethnic groups are shown in Table 1. Black and Asian ethnicity patients were significantly younger with a median age of 58 years (Asian) and 60 years (Black), compared with 70 years in the White group (p<0.001). Comorbidity data were available in 5518 (99.7%) of patients and burden of comorbid disease varied between ethnic groups in prevalence, type, and age distribution. Diabetes and CKD were more prevalent at an earlier age in Asian and Black patients, frailty and dementia were more prevalent in older White patients (Table 1, Table S2). UK National Early Warning Score within the first day of admission was clinically similar between groups with a 0.5 difference in mean scores between the highest and lowest groups (2.8 in Mixed/Other, 2.3 in Black). Around one in five patients developed early acute kidney injury (AKI) within seven days of hospital admission, rates of AKI were highest in the Black group (25%). Peak C-reactive protein (CRP) during admission did not differ significantly between groups however peak D-dimer was higher in the Black population (median 2.23 mg/L) compared with other ethnicities (median 1.21 mg/L Asian, 1.54 mg/L White).

**Table 1:**
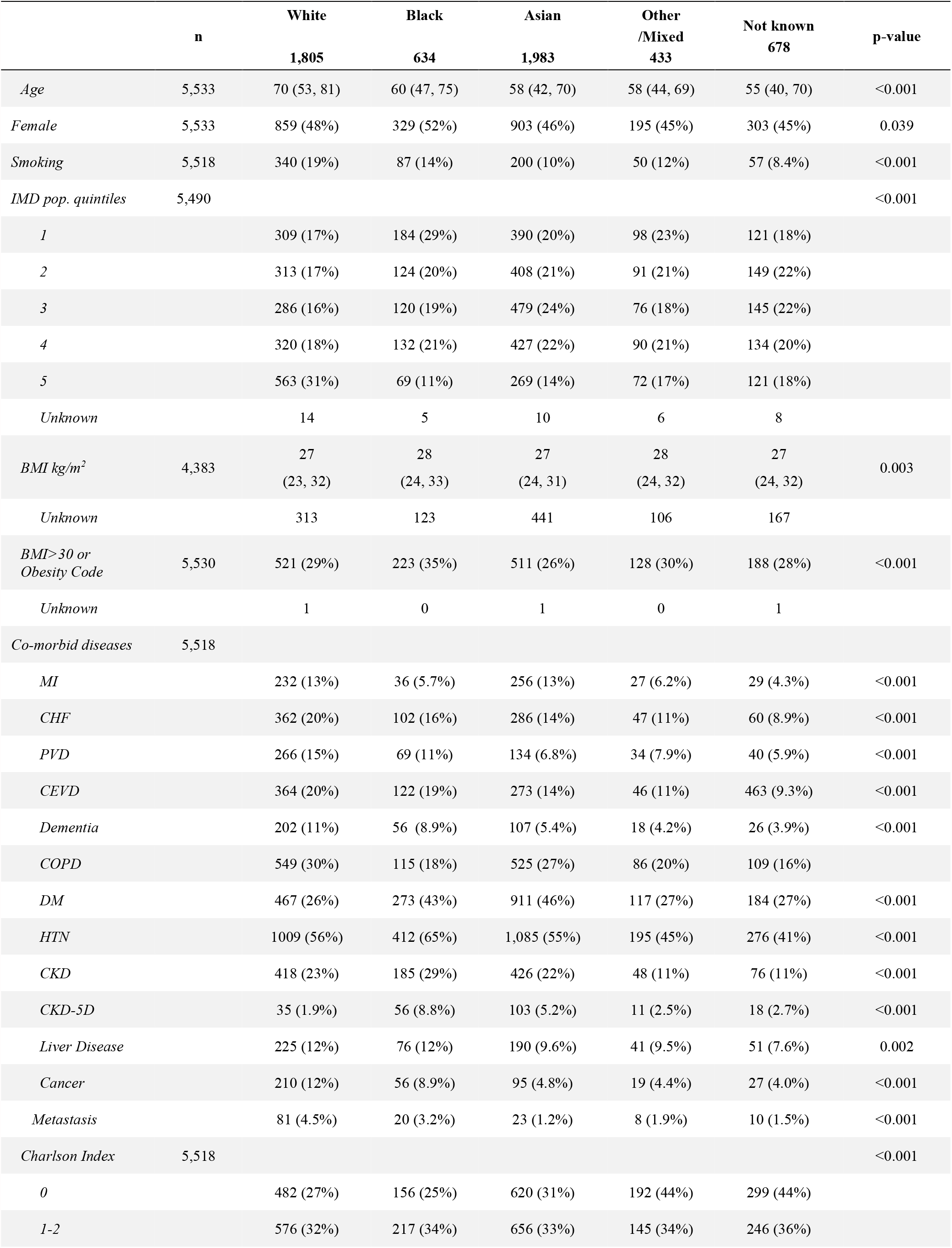
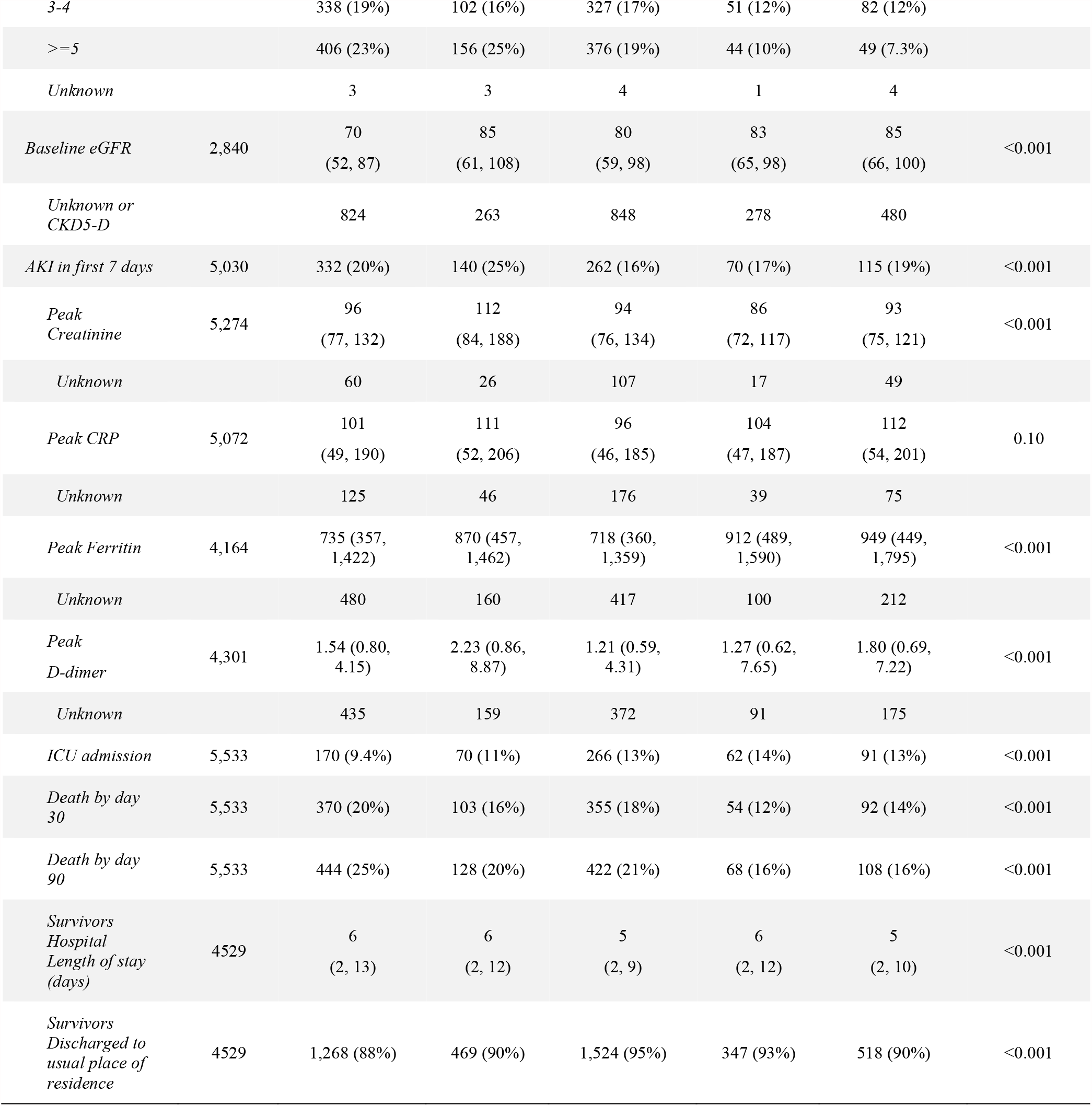
Characteristics of 5533 patients with COVID-19 associated first hospital admissions during second pandemic wave between 1/9/20 and 17/2/21.

|  | **n** | **White**<br>**1,805** | **Black**<br>**634** | **Asian**<br>**1,983** | **Other**<br>**/Mixed**<br>**433** | **Not known**<br>**678** | **p-value** |
| --- | --- | --- | --- | --- | --- | --- | --- |
| *Age* | 5,533 | 70 (53, 81) | 60 (47, 75) | 58 (42, 70) | 58 (44, 69) | 55 (40, 70) | <0.001 |
| *Female* | 5,533 | 859 (48%) | 329 (52%) | 903 (46%) | 195 (45%) | 303 (45%) | 0.039 |
| *Smoking* | 5,518 | 340 (19%) | 87 (14%) | 200 (10%) | 50 (12%) | 57 (8.4%) | <0.001 |
| *IMD pop. quintiles* | 5,490 |  |  |  |  |  | <0.001 |
| *1* |  | 309 (17%) | 184 (29%) | 390 (20%) | 98 (23%) | 121 (18%) |  |
| *2* |  | 313 (17%) | 124 (20%) | 408 (21%) | 91 (21%) | 149 (22%) |  |
| *3* |  | 286 (16%) | 120 (19%) | 479 (24%) | 76 (18%) | 145 (22%) |  |
| *4* |  | 320 (18%) | 132 (21%) | 427 (22%) | 90 (21%) | 134 (20%) |  |
| *5* |  | 563 (31%) | 69 (11%) | 269 (14%) | 72 (17%) | 121 (18%) |  |
| *Unknown* |  | 14 | 5 | 10 | 6 | 8 |  |
| *BMI kg/m<sup>2</sup>* | 4,383 | 27<br>(23, 32) | 28<br>(24, 33) | 27<br>(24, 31) | 28<br>(24, 32) | 27<br>(24, 32) | 0.003 |
| *Unknown* |  | 313 | 123 | 441 | 106 | 167 |  |
| *BMI> 30 or*<br>*Obesity Code* | 5,530 | 521 (29%) | 223 (35%) | 511 (26%) | 128 (30%) | 188 (28%) | <0.001 |
| *Unknown* |  | 1 | 0 | 1 | 0 | 1 |  |
| *Co-morbid diseases* | 5,518 |  |  |  |  |  |  |
| *MI* |  | 232 (13%) | 36 (5.7%) | 256 (13%) | 27 (6.2%) | 29 (4.3%) | <0.001 |
| *CHF* |  | 362 (20%) | 102 (16%) | 286 (14%) | 47 (11%) | 60 (8.9%) | <0.001 |
| *PVD* |  | 266 (15%) | 69 (11%) | 134 (6.8%) | 34 (7.9%) | 40 (5.9%) | <0.001 |
| *CEVD* |  | 364 (20%) | 122 (19%) | 273 (14%) | 46 (11%) | 463 (9.3%) | <0.001 |
| *Dementia* |  | 202 (11%) | 56 (8.9%) | 107 (5.4%) | 18 (4.2%) | 26 (3.9%) | <0.001 |
| *COPD* |  | 549 (30%) | 115 (18%) | 525 (27%) | 86 (20%) | 109 (16%) |  |
| *DM* |  | 467 (26%) | 273 (43%) | 911 (46%) | 117 (27%) | 184 (27%) | <0.001 |
| *HTN* |  | 1009 (56%) | 412 (65%) | 1,085 (55%) | 195 (45%) | 276 (41%) | <0.001 |
| *CKD* |  | 418 (23%) | 185 (29%) | 426 (22%) | 48 (11%) | 76 (11%) | <0.001 |
| *CKD-5D* |  | 35 (1.9%) | 56 (8.8%) | 103 (5.2%) | 11 (2.5%) | 18 (2.7%) | <0.001 |
| *Liver Disease* |  | 225 (12%) | 76 (12%) | 190 (9.6%) | 41 (9.5%) | 51 (7.6%) | 0.002 |
| *Cancer* |  | 210 (12%) | 56 (8.9%) | 95 (4.8%) | 19 (4.4%) | 27 (4.0%) | <0.001 |
| *Metastasis* |  | 81 (4.5%) | 20 (3.2%) | 23 (1.2%) | 8 (1.9%) | 10 (1.5%) | <0.001 |
| *Charlson Index* | 5,518 |  |  |  |  |  | <0.001 |
| *0* |  | 482 (27%) | 156 (25%) | 620 (31%) | 192 (44%) | 299 (44%) |  |
| *1-2* |  | 576 (32%) | 217 (34%) | 656 (33%) | 145 (34%) | 246 (36%) |  |

|  |  |  |  |  |  |  |  |  |
| --- | --- | --- | --- | --- | --- | --- | --- | --- |
|  |  | 3-4 | 338 (19%) | 102 (16%) | 327 (17%) | 51 (12%) | 82 (12%) |  |
|  |  | >=5 | 406 (23%) | 156 (25%) | 376 (19%) | 44 (10%) | 49 (7.3%) |  |
|  |  | Unknown | 3 | 3 | 4 | 1 | 4 |  |
| Baseline eGFR | 2,840 |  | 70<br>(52, 87) | 85<br>(61, 108) | 80<br>(59, 98) | 83<br>(65, 98) | 85<br>(66, 100) | <0.001 |
|  |  | Unknown or<br>CKD5-D | 824 | 263 | 848 | 278 | 480 |  |
| AKI in first 7 days | 5,030 |  | 332 (20%) | 140 (25%) | 262 (16%) | 70 (17%) | 115 (19%) | <0.001 |
| Peak<br>Creatinine | 5,274 |  | 96<br>(77, 132) | 112<br>(84, 188) | 94<br>(76, 134) | 86<br>(72, 117) | 93<br>(75, 121) | <0.001 |
|  |  | Unknown | 60 | 26 | 107 | 17 | 49 |  |
| Peak CRP | 5,072 |  | 101<br>(49, 190) | 111<br>(52, 206) | 96<br>(46, 185) | 104<br>(47, 187) | 112<br>(54, 201) | 0.10 |
|  |  | Unknown | 125 | 46 | 176 | 39 | 75 |  |
| Peak Ferritin | 4,164 |  | 735 (357,<br>1,422) | 870 (457,<br>1,462) | 718 (360,<br>1,359) | 912 (489,<br>1,590) | 949 (449,<br>1,795) | <0.001 |
|  |  | Unknown | 480 | 160 | 417 | 100 | 212 |  |
| Peak<br>D-dimer | 4,301 |  | 1.54 (0.80,<br>4.15) | 2.23 (0.86,<br>8.87) | 1.21 (0.59,<br>4.31) | 1.27 (0.62,<br>7.65) | 1.80 (0.69,<br>7.22) | <0.001 |
|  |  | Unknown | 435 | 159 | 372 | 91 | 175 |  |
| ICU admission | 5,533 |  | 170 (9.4%) | 70 (11%) | 266 (13%) | 62 (14%) | 91 (13%) | <0.001 |
| Death by day<br>30 | 5,533 |  | 370 (20%) | 103 (16%) | 355 (18%) | 54 (12%) | 92 (14%) | <0.001 |
| Death by day<br>90 | 5,533 |  | 444 (25%) | 128 (20%) | 422 (21%) | 68 (16%) | 108 (16%) | <0.001 |
| Survivors<br>Hospital<br>Length of stay<br>(days) | 4529 |  | 6<br>(2, 13) | 6<br>(2, 12) | 5<br>(2, 9) | 6<br>(2, 12) | 5<br>(2, 10) | <0.001 |
| Survivors<br>Discharged to<br>usual place of<br>residence | 4529 |  | 1,268 (88%) | 469 (90%) | 1,524 (95%) | 347 (93%) | 518 (90%) | <0.001 |

### Age- and sex-adjusted survival

Overall raw 30-day mortality was 17.6%, ranging from 20% in the White group to 12% in the Mixed/Other group. We considered 4855 patients with documented ethnicity in the primary outcome analysis, of these 46 (0.95%) had incomplete data and were excluded from multivariable survival analysis. After adjustment for between-group differences in age, sex, presence of diabetes, CKD, hypertension, smoking history, obesity, and socioeconomic deprivation, patients with Asian ethnicity were at significantly higher risk of death within 30 days compared to White patients (HR 1.47 [1.24-1.73]). No association with increased risk of death was observed in the Black or Mixed/Other groups (Table 2, Fig 1a). There was no statistical evidence for violation of the proportional-hazards assumption. Increased rate of death in the Asian group was also demonstrated in analyses including total CCI (HR 1.47 [1.26-1.72]) or HFRS (HR 1.62 [1.38-1.89]) rather than specific comorbidities (Table 2, Figs S3-4). This effect was persistent when the initial analysis was extended to 90-day mortality (HR for Asian ethnicity 1.41 [1.22-1.64]) (Table 2, Figs 1b and 2).

**Table 2:** Multivariable modelling (PH: Proportional Hazards, LR: Logistic Regression).

|  | *Second wave* |  |  |  |  | *First wave* |
| --- | --- | --- | --- | --- | --- | --- |
| *Outcome* | 30-day mortality | 30-day mortality | 30-day mortality | 90-day mortality | ICU admission | 1-year survival |
| *Model* | PH | PH | PH | PH | LR | PH |
| *Number* | 4809 | 4809 | 4809 | 4809 | 4809 | 1722 |
| *Events* | 879 | 879 | 879 | 1057 | 459 | 586 |
| *Age*<br>(75<sup>th</sup> vs. 25<sup>th</sup> centiles) | 5.85<br>(4.93-6.94) | 5.12<br>(4.33-6.06) | 5.09<br>(4.29-6.02) | 5.22<br>(4.48-6.08) | 0.70<br>(0.58-0.84) | 4.33<br>(3.58-5.24) |
| *Male Sex* | 1.44<br>(1.26-1.66) | 1.50<br>(1.31-1.73) | 1.43<br>(1.25-1.67) | 1.38<br>(1.22-1.57) | 1.87<br>(1.54-2.26) | 1.42<br>(1.19-1.68) |
| *Ethnicity* |  |  |  |  |  |  |
| *White* | *Ref* | *Ref* | *Ref* | *Ref* | *Ref* | *Ref* |
| *Asian* | 1.47<br>(1.24-1.73) | 1.62<br>(1.38-1.89) | 1.47<br>(1.26-1.72) | 1.41<br>(1.22-1.64) | 1.38<br>(1.10-1.73) | 1.38<br>(1.12-1.71) |
| *Black* | 1.06<br>(0.84-1.33) | 1.16<br>(0.92-1.45) | 1.06<br>(0.85-1.33) | 1.07<br>(0.87-1.31) | 1.09<br>(0.80-1.48) | 1.14<br>(0.90-1.44) |
| *Mixed/Other* | 1.03<br>(0.77-1.39) | 1.09<br>(0.81-1.46) | 1.10<br>(0.82-1.47) | 1.06<br>(0.82-1.38) | 1.53<br>(1.11-2.11) | 1.05<br>(0.74-1.48) |
| *Smoking* | 1.23<br>(1.03-1.47) | 1.14<br>(0.95-1.36) | 1.14<br>(0.95-1.36) | 1.19<br>(1.01-1.41) | 0.90<br>(0.69-1.18) | 1.20<br>(0.95-1.51) |
| *Obesity* | 1.36<br>(1.17-1.61) | 1.44<br>(1.23-1.69) | 1.37<br>(1.17-1.61) | 1.27<br>(1.10-1.47) | 1.63<br>(1.35-1.98) | 1.16<br>(0.97-1.40) |
| *Diabetes* | 1.09<br>(0.94-1.26) |  |  | 1.01<br>(0.89-1.16) | 1.13<br>(0.92-1.39) | 1.28<br>(1.07-1.53) |
| *CKD* | 1.34<br>(1.16-1.55) |  |  | 1.44<br>(1.26-1.64) | 0.95<br>(0.75-1.21) | 1.39<br>(1.16-1.66) |
| *Hypertension* | 1.24<br>(1.03-1.49) |  |  | 1.28<br>(1.08-1.51) | 1.46<br>(1.14-1.87) | 0.98<br>(0.78-1.23) |
| *CCI 0* |  |  | *Ref* |  |  |  |
| *CCI 1-2* |  |  | 2.24<br>(1.66-3.02) |  |  |  |
| *CCI 3-4* |  |  | 2.78<br>(2.04-3.81) |  |  |  |
| *CCI >5* |  |  | 3.62<br>(2.67-4.91) |  |  |  |
| *HFRS <5* |  | *Ref* |  |  |  |  |
| *HFRS 5-15* |  | 2.29<br>(1.94-2.70) |  |  |  |  |
| *HFRS >15* |  | 2.10<br>(1.70-2.60) |  |  |  |  |
| *IMD 1* | *Ref* | *Ref* | *Ref* | *Ref* | *Ref* | *Ref* |
| *IMD 2* | 0.87<br>(0.70-1.07) | 0.84<br>(0.68-1.04) | 0.85<br>(0.69-1.05) | 0.93<br>(0.77-1.13) | 1.0<br>(0.76-1.32) | 0.97<br>(0.75-1.27) |
| *IMD 3* | 0.77<br>(0.62-0.95) | 0.77<br>(0.62-0.96) | 0.77<br>(0.62-0.95) | 0.81<br>(0.67-0.98) | 0.89<br>(0.67-1.17) | 0.87<br>(0.67-1.14) |
| *IMD 4* | 1.01<br>(0.82-1.24) | 1.00<br>(0.82-1.23) | 1.00<br>(0.82-1.23) | 1.05<br>(0.87-1.27) | 0.76<br>(0.57-1.01) | 0.93<br>(0.72-1.20) |
| *IMD 5* | 0.92<br>(0.75-1.13) | 0.92<br>(0.74-1.13) | 0.91<br>(0.74-1.12) | 0.90<br>(0.75-1.10) | 1.00<br>(0.74-1.32) | 1.05<br>(0.81-1.35) |

**Fig 1:**
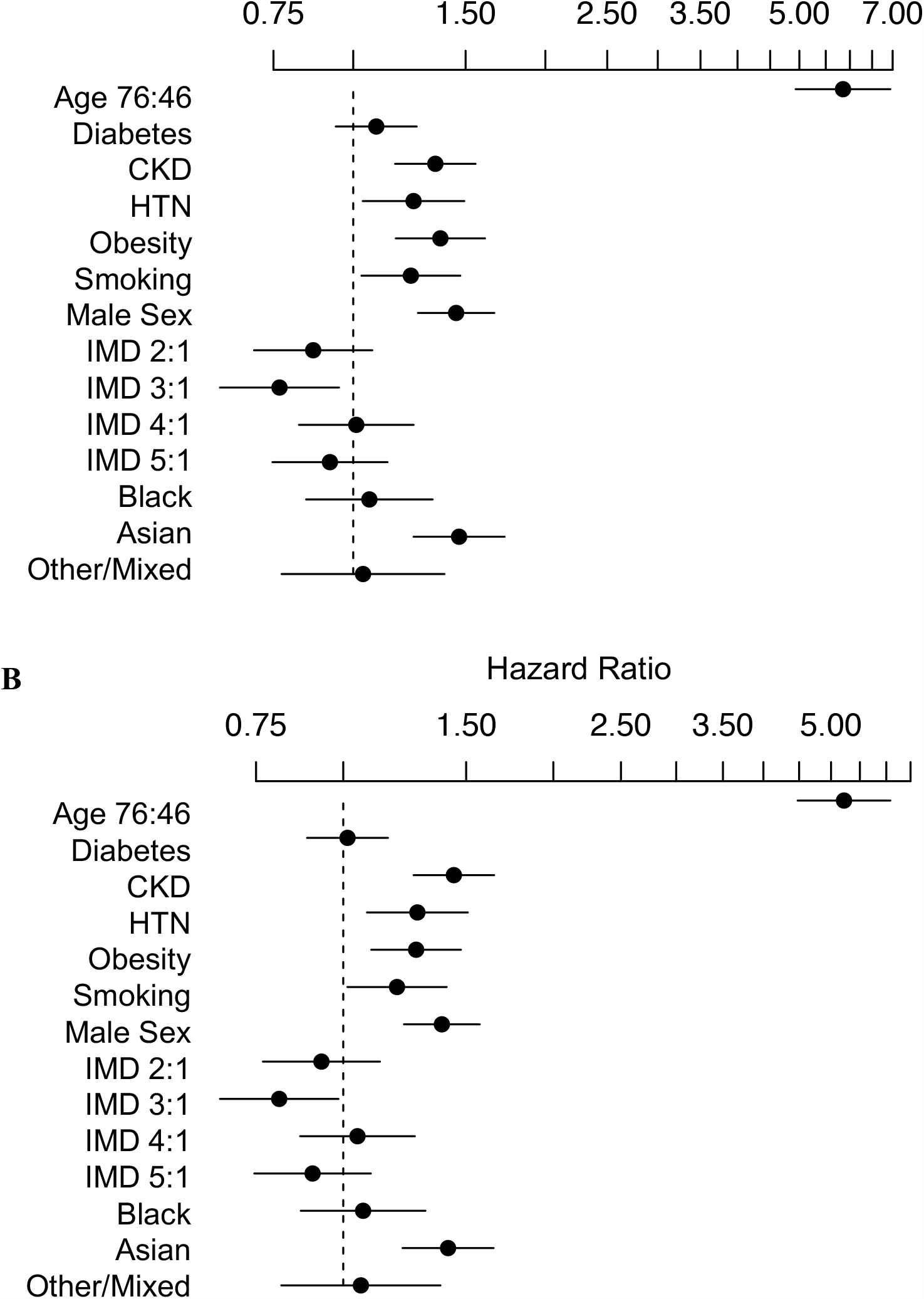
Cox-proportional Hazard analysis included covariates: Age, Sex, Hypertension, Diabetes, Chronic Kidney Disease, Smoking history, Obesity, IMD (quintiles of local study population) for survival over 30 (A) and 90 (B) days in the second pandemic wave *ETHICAL* cohort.

**Fig 2:**
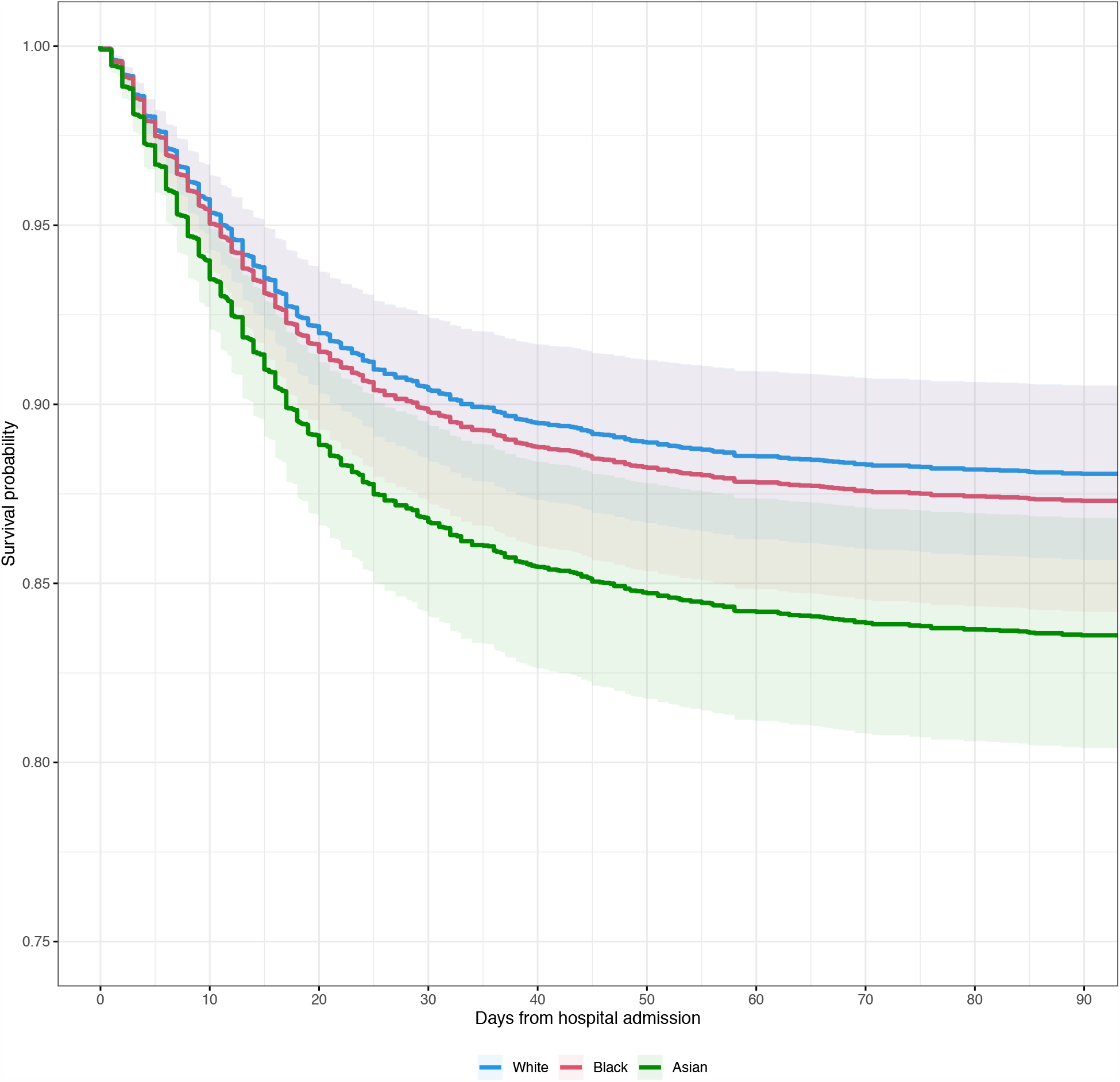
Predicted survival over 90-days by Ethnicity in a 65-year-old Male living in IMD-3 without Diabetes, CKD, Smoking History, Hypertension or Obesity. Based on second wave Cox-proportional Hazard analysis included covariates: Age, Sex, Hypertension, Diabetes, Chronic Kidney Disease, Smoking history, Obesity, IMD (quintiles of local study population).

### ICU admission

ICU admission occurred in 11.9% of cases and was lowest in the White group (9.4%) and highest in the Asian (13%) and Mixed/Other groups (14%) (Table 1). In multivariable analysis excluding unknown ethnicity and unknown comorbidity (n=4809), Asian (OR 1.38 [1.10-1.73]) and Mixed/Other (OR 1.53 [1.11-2.11]) ethnicities were associated with increased age-, sex-, and comorbidity-adjusted risk of ICU admission (Table 2, Fig S5).

### Extended follow up of first wave patients

All 1996 patients admitted during the first wave were re-analysed 12 months after admission. Overall, 365-day mortality was 32.9% with only a further 2.4% of the initial population dying between days 90 and 365. Age-, sex-, and comorbidity-adjusted survival analysis in 1722 patients with known ethnicity and comorbidity data is shown in Fig 3. In this analysis adjusted risk of death associated with Asian ethnicity remained significant (HR 1.38 [1.12-1.71]) (Table 2, Fig S6). However, the trend we previously observed for increased mortality in Black patients was not sustained over time. In this analysis, ethnicity showed significant violation of the proportional-hazards assumption. Remodelling with stratification by ethnicity (Fig 3) suggested sustained, proportional increases in risk of death in Asian patients, but that the initially higher rate of death in Black patients was attenuated over time so that predicted survival at one-year was similar to White patients, after controlling for age, sex and other risk factors.

**Fig 3:**
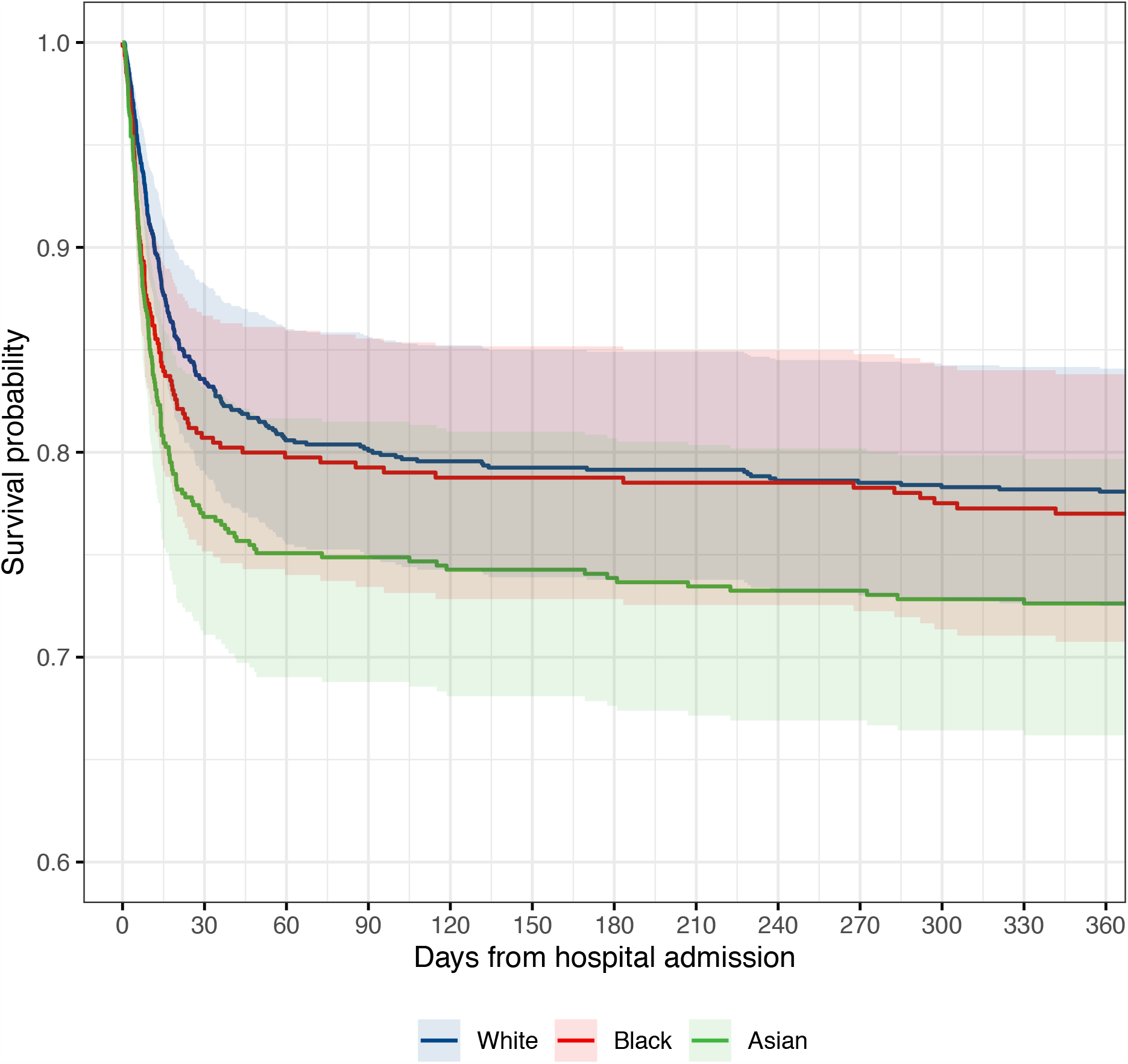
Predicted survival by Ethnicity in a 65-year-old Male living in IMD-3 without Diabetes, CKD, Smoking History, Hypertension or Obesity based on 12-month follow-up of the first pandemic wave *ETHICAL* population in a Cox-proportional Hazard analysis stratified by ethnicity. Included covariates: Age, Sex, Hypertension, Diabetes, Chronic Kidney Disease, Smoking history, Obesity, IMD (quintiles of local study population). Excess mortality associated with Asian ethnicity has persisted over follow up while early survival disadvantage associated with Black ethnicity has attenuated with longer follow-up. Other/Mixed group omitted for clarity.

### Comparison of second and first waves

Overall, in comparison with the first wave data, significantly more patients were included (5533 vs 1996). Median age was lower in the second wave in White (70 vs 73, p<0.001), Black (60 vs 64, p<0.001) but not Asian (58 vs 59, p=0.21) groups. In the second wave, a larger proportion of admissions were in Asian patients (35.7% vs. 27.0%, p<0.001) and a smaller proportion in Black patients (11.4% vs 17.0%, p<0.001). While the proportion of White patients in the second wave also decreased, this change was relatively slight (32.6% vs 35.8%, p=0.04). Overall raw 30-day mortality was 17.6% compared to 27.4% in the first wave cohort (p<0.001). Furthermore, predicted age-, sex-, and comorbidity-adjusted survival improved in all groups: predicted 30-day mortality in a 65-year-old male of White ethnicity without diabetes, CKD, hypertension, obesity, smoking history, and median IMD was 9.0% (6.9-11.1%) in the second wave model, compared to 16.4% (11.8-20.9%) in the first wave model (Figs 2, 3). There was a similar difference across ethnic groups, for a similar patient of Asian ethnicity, predicted 30-day mortality was 12.9% (10.0-15.7%) in the second wave versus 23.2% (16.9-28.9%) in the first (Figs 2, 3). Other notable changes included lower peak CRPs in the second wave across all groups and lower rates of ICU admission both overall (11.9 vs 15.9%, p<0.001) and within ethnic groups.

### Admission rates

Admissions within each ethnic group by age group and the local population distribution are shown in Tables S3-6 and Figure 4. Relative to the White population, local Asian and Black populations experienced between two to four-fold higher age-standardised rates of hospital admissions during the COVID-19 first and second waves, with a 4.07 (3.77-4.39) times higher rate in Asian patients in the second wave (Table 3). By comparison during 2013-18, age-standardised rates of emergency admissions in local Black and Asian populations were only 10-30% greater than in White local-residents. Finally, up to 10% of hospital admissions were recorded as of unknown ethnicity, a categorisation not used in the 2011 UK census.

**Table 3:** Hospital admission rates per 100,000 of the age-standardized local population by ethnic group (based on European standard population for ages 15 to 85+).

|  | *Admissions/100,000*<br>*Age-Standardized Population* | *95%CI* | *Ratio vs. White*<br>*Ethnicity* | *95% CI* |
| --- | --- | --- | --- | --- |
| **Wave 1** |  |  |  |  |
| *White* | 239 | 217-260 | 1.00 |  |
| *Black* | 727 | 641-814 | **3.05** | **2.61-3.52** |
| *Asian* | 584 | 529-639 | **2.45** | **2.37-3.04** |
| *Mixed* | 225 | 92-358 | 0.94 | 0.38-1.51 |
| *Other* | 898 | 715-1082 | **3.76** | **2.94-4.63** |
| **Wave 2** |  |  |  |  |
| *White* | 532 | 500-563 | 1.00 |  |
| *Black* | 1,116 | 1017-1216 | **2.10** | **1.88-2.33** |
| *Asian* | 2,163 | 2058-2267 | **4.07** | **3.77-4.39** |
| *Mixed* | 451 | 295-607 | 0.85 | 0.55-1.15 |
| *Other* | 2,223 | 1947-2499 | **4.18** | **3.62-4.77** |
| **2013-18** |  |  |  |  |
| *White* | 20,056 | 19875-20236 | 1.00 |  |
| *Black* | 22,403 | 22021-22785 | **1.12** | **1.1-1.14** |
| *Asian* | 26,823 | 26522-27124 | **1.34** | **1.32-1.36** |
| *Mixed* | 13,265 | 12633-13896 | **0.66** | **0.63-0.69** |
| *Other* | 36,300 | 35381-37220 | **1.81** | **1.76-1.86** |

**Fig 4:**
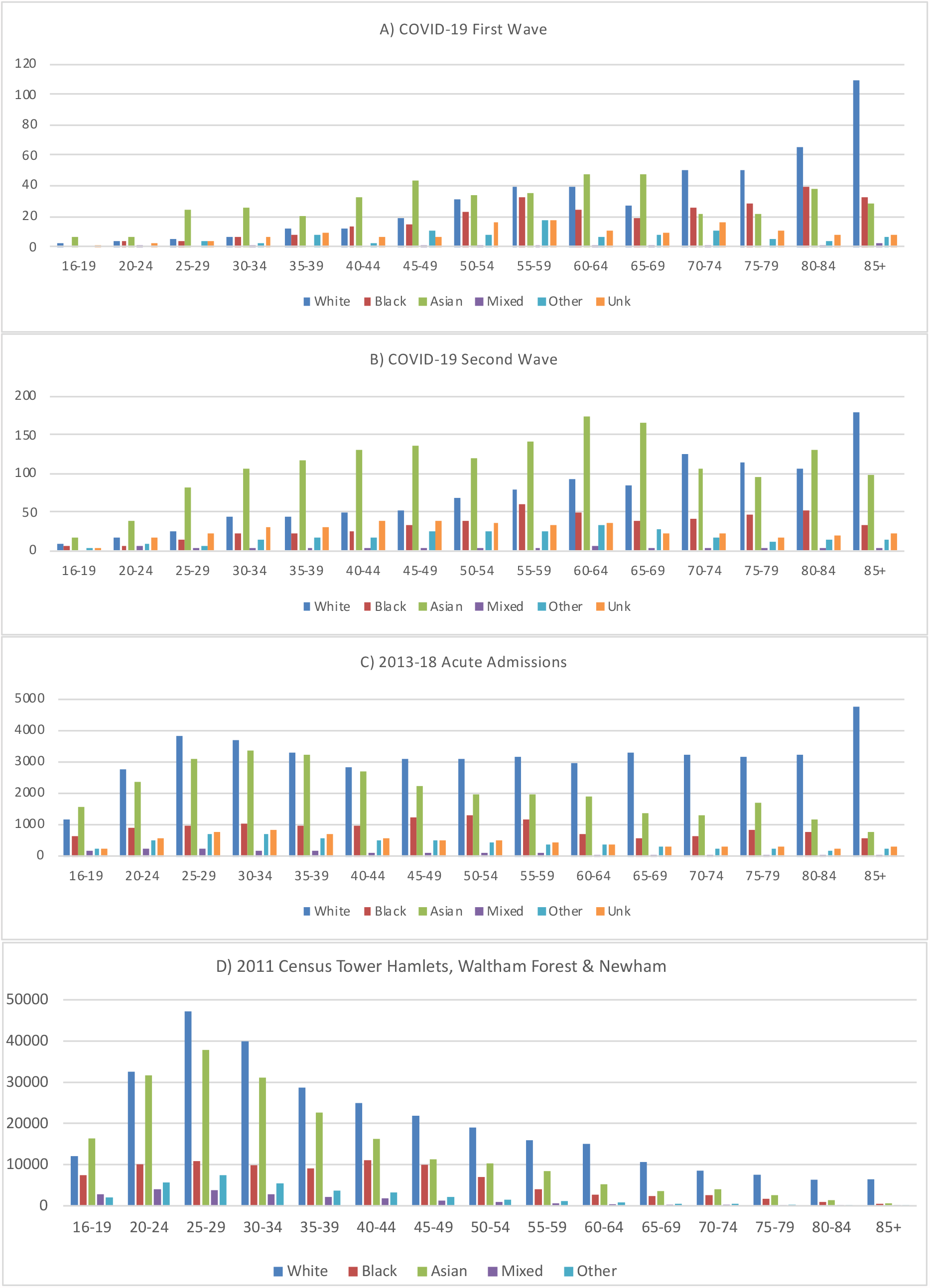
Numbers of first admissions to Barts Health hospitals in residents of the London Boroughs of Tower Hamlets, Newham, and Waltham Forest aged ≥16, grouped by age and ethnicity for the first and second waves of COVID-19 (panels A & B) or any acute admissions during 2013-18 (panel C). Age and Ethnicity distribution of the Tower Hamlets, Newham, and Waltham Forest population in the 2011 UK Census is shown for comparison (panel D).

## DISCUSSION

The principal finding of this study was that amongst 5533 patients hospitalised with COVID-19 in the ethnically diverse area of east London during the second wave, there remained important ethnic disparities in patient outcomes. In the second wave, the proportion of patient-admissions of Asian ethnicity increased, most markedly in the younger age groups. In the second wave the increased risk of death amongst Asian patients seen in the first wave persisted, whilst patients of Black ethnicity had comparable outcomes to White patients in the second wave. Similarly, the increased risk of death in Asian patients admitted during the first wave continued during prolonged follow-up whereas Black patients showed no longer-term risk compared to White inpatients.

We considered differential rates of admission between ethnic groups as an additional contributor to ethnic imbalances in the impact of COVID-19. By comparing age and ethnicity from inpatients drawn from three boroughs in east London with the local population, we were able to demonstrate that rates of admission were 2-4-fold higher in Black and Asian compared to White populations with a similar age distribution. This effect particularly prominent in the Asian population in the second wave and the Black population in the first. Importantly, these increased rates of admission in ethnic minority groups were substantially higher than seen in emergency admissions to Barts Health hospitals from the same local community prior to COVID. Notably, in contrast to our findings in COVID-19, in the pre-COVID cohort Black and Asian ethnicities had lower age-, sex-, and comorbidity-adjusted risk of death compared to white emergency admissions (manuscript under review), similar to findings for non-COVID death reported from the OPENSafely platform.^17^ Furthermore, a significant number of admissions with unknown ethnicity and a small excess of patients classified as of Other ethnicity are likely to reflect a substantial number of additional patients with non-White background, if anything increasing the excess rate of admissions in minority categories.

Compared to the first wave, mortality was over one third lower across all ethnic groups, even after adjustment for age and comorbidity differences. This may indicate improvements in specific treatments,^18,19^ including routine early use of corticosteroids,^20^ and in processes of care,^21^ including the development of purpose built a 150-bed COVID-ICU at the Royal London Hospital.^22^ In addition, the total patient population appeared less acutely unwell with fewer ICU admissions, lower levels of AKI, and lower peak markers of inflammation. This could reflect treatment changes both in and before hospitalisation or a change in disease profile over time. However, despite apparent improvements in outcomes overall excess risk of death in patients of Asian background persisted in both the second wave and prolonged follow-up of the first. In contrast, early risk of death in Black patients in the first wave was not sustained over prolonged follow-up, and in the second wave, risk of death following hospital admission associated with Black ethnicity did not differ significantly to White ethnicity. This finding is in line with other studies examining the first wave data in hospitalised patients across the UK^23^ and early second wave data examining COVID-19 deaths in a large primary care dataset.^24^ Importantly, our results suggest that differential rates of hospitalisation rather than hospital outcomes themselves may be the strongest ethnic inequality driving adverse outcomes associated with COVID-19, particularly for Black patients. This suggests that COVID acquisition, rather than delayed presentation or response to treatment in hospital is one of the leading drivers of excess mortality in these populations. Those of South Asian background do appear doubly affected experiencing an almost 4-fold higher age-matched rate of hospitalisation as well as a 30-40% increased chance of death once hospitalised compared to similar White patients. Importantly, these differences have persisted despite improvement in treatments, processes of care, and COVID-19 outcomes in general.

### Reasons for ethnicity inequality in COVID outcomes

Disparities in health outcomes and mortality rates between people of different ethnicities has been the subject of intense public and medical scrutiny and debate. Numerous potential drivers for these inequalities have been suggested including differences in socioeconomic status, occupational exposure, housing density, access to home working, multi-generational living, engagement with and access to healthcare services, incidence and severity of comorbid disease.^25,26^ Furthermore, the effects of racism and structural discrimination are an important underpinning consideration.^27^ Although we have attempted to account for some of these factors such as comorbidity differences, presence or absence of conditions such as diabetes of hypertension may fail to account for differences in disease severity and duration of exposure between ethnic groups. Similarly, postcode-based assessment of deprivation may fail to account for differences in the relative poverty between ethnic groups living in the same community. Notably, this study population is drawn from a community with a relatively high level of deprivation with comparatively little variation, which may account for a lack of association between deprivation and survival in our analysis. Consequently, the White comparator population in this study may represent a group with worse underlying health than the general UK population.

### Strengths and limitations

This analysis supports the findings of several published and pre-printed analyses examining ethnicity and COVID-19 outcomes in the UK but is distinguished by the large representation of ethnic minority groups and large absolute numbers of these patients drawn from a single geographic region and treated within the same hospital system. Importantly previous analyses are drawn from large populations which may poorly represent the most ethnically diverse areas of the UK. The OPENSAFELY dataset was only 6% Asian and 2% Black in the first wave,^3^ and 7.2% Asian 1% Black in the second wave,^28^ the Second Generation Surveillance System (SGSS)/COVID-19 Hospitalization in England Surveillance System (CHESS) dataset 8.4% Asian 3.8% Black,^29^ and the ISARIC dataset 4.5% South Asian and 3.6% Black.^23^ In terms of absolute patients numbers, this study thus represents one of the largest descriptor of outcomes in minority ethnic patients with COVID-19 (Table S7) and is strengthened by comparison to a White population drawn from the same location experiencing the same treatment, eliminating many of the geographic biases present within larger studies with poorer minority ethnic representation. Our analyses are strengthened by adherence to a prespecified analysis plan, inclusion of a range of baseline, comorbidity, and COVID-19 risk factors in multivariable modelling, and sensitivity tests using different measures of comorbidity. However as with all observational studies, not all potential contributing risk factors could be assessed, including other measures of baseline health status such as nutritional or lifestyle influences. We were also unable to compare the effect of the differing viral strains in the first and second wave. Other limitations include not being able to include suspected but not proven COVID-19 cases or community cases not requiring hospital admission. In addition, contextual factors such as household composition and occupation could not be assessed. Currently, ethnic categorisations used in healthcare do not reflect the vast heterogeneity within each aggregated ethnic category. There may be specific differences in presentation and outcomes particularly across Black people (Black African versus Black Caribbean) and Asian people (Bangladeshi/Pakistani versus Indian). Similarly, using a composite measure to assess socioeconomic deprivation limits the ability to evaluate varying effects of sub-domains. Finally, as previously discussed, our study population has a particularly high relative level of socioeconomic deprivation overall and the variation in patterns of deprivation between and within ethnic groups may differ with other study cohorts.

### Conclusion

Despite improvement in overall outcomes, better treatments, and processes of care, Asian and Black ethnic groups continue to have an increased hospital admission and death in hospital associated with COVID-19. Our findings suggest that COVID acquisition rather than delayed presentation or response to treatment in hospital may be one of the leading drivers of excess mortality in the ethnic minority populations in east London. This confirms the need to understand and contextualise structural and socioeconomic factors that drive these outcomes. To achieve this, qualitative data will need to be aligned with insight from community-based qualitative research exploring the lived realities of minority ethnic communities in order to inform the implementation of impactful community-level interventions.

## Data Availability

The statistical analysis plan can be accessed online. The authors will be happy to consider additional analyses of the anonymised dataset on request. The need for stringent measures to prevent re-identification of individuals within a discrete geographical location and limited time-period however preclude sharing of patient level dataset in a GDPR compliant form.

## Contributions

Study Concept and Design YIW VJA CMO RD RMP ZP JRP

Ethics application and Approvals VJA JRP

Study protocol and analysis plan YIW JRP

Data Extraction YIW JRP

Data Analysis YIW JRP

Critical review of finding All authors

Manuscript writing YIW JRP

Review of final submission All authors

## Ethics approval

This study was approved by NHS England Health Research Authority and Yorkshire & The Humber - Bradford Leeds Research Ethics Committee and approved as anonymised analysis of routinely collected patient data without need for direct consent (Ethics reference 20/YH/0159).

## Funding

This research received no specific grant from any funding agency in the public, commercial or not-for-profit sectors.

## Declarations of interest

All authors declare no other competing interests.

## Notes

### Competing Interest Statement

The authors have declared no competing interest.

