## Supplement for "Ethnic disparities in hospitalisation and hospital-outcomes during the second wave of COVID-19 infection in east London"

### Supplementary material

#### Definition of key variables

*Ethnicity*
We defined ethnic groups using the 16+1 categories defined in the 2001 census which form the UK national mandatory standard for the collection and analysis of ethnicity in the NHS data dictionary. Importantly, in the UK 'Asian' ethnic category refers predominantly to those of a South Asian background (including Indian, Pakistani and Bangladeshi), while patients of a Chinese background are placed in the 'Other Ethnic Groups' category.

| | |
|---|---|
| White | A British<br>B Irish<br>C Any other White background |
| Mixed | D White and Black Caribbean<br>E White and Black African<br>F White and Asian<br>G Any other mixed background |
| Asian or Asian British | H Indian<br>J Pakistani<br>K Bangladeshi<br>L Any other Asian background |
| Black or Black British | M Caribbean<br>N African<br>P Any other Black background |
| Other Ethnic Groups | R Chinese<br>S Any other ethnic group |
| +1 category | Z Not stated (Reserved for cases where patients declined to provide information) |

In order to preserve statistical power to detect differences between groups, pre-specified analysis was carried out between ethnicity defined by the 5-high level groups White, Mixed, Asian or Asian British, Black or Black British and Other with merging of the "Mixed" and "Other" categories. Category Z was excluded from our primary analysis as were cases where no ethnicity data was recorded (Unknown).

*Index of Multiple Deprivation*
Index of Multiple Deprivation (IMD) was defined from patient home address postcode using UK government statistics (https://www.gov.uk/government/statistics/english-indices-of-deprivation-2019). Matching of Lower-layer Super Output Areas (LSOAs) was undertaken against the Office of National Statistics Postcode Directory (ONSPD) February 2020 datafile (https://geoportal.statistics.gov.uk/datasets/ons-postcode-directory-february-2020: accessed on 1st May 2020). IMD was presented as quintiles within England using raw scores for descriptive results and quintiles within the study cohort in multivariable analysis.

*Smoking*
History of tobacco use was defined by presence of the WHO ICD-10 codes F17·1-F17·2, Z72·0, Z87·8, Z71·6 and T65·2.

*Comorbidity*
Diagnosis of co-morbidities and assignment of Charlson Comorbidity Index was based on mapping from ICD-10 coding from previous admissions using the mapping of Quan H, et al.
*Quan H, et al. Coding algorithms for defining comorbidities in ICD-9-CM and ICD-10 administrative data. Med Care 2005;43(11):1130-9.*
Diagnosis of Hypertension was based on mapping ICD-10 codes to the Elixhauser comorbidity index.
*Elixhauser A, et al. Comorbidity measures for use with administrative data. Med Care 1998;36:8-27.*

*Hospital frailty risk score*
Hospital frailty risk score was calculated from mapping ICD-10 coding of hospital attendances.
*Gilbert T, et al. Development and validation of a Hospital Frailty Risk Score focusing on older people in acute care settings using electronic hospital records: an observational study. Lancet 2018;391(10132):1775-1782.*

*Acute Kidney injury*
Acute kidney injury (AKI) within first 7 days of admission was defined using the KDIGO 2012 creatinine criteria either a 1·5-fold rise over baseline within 7 days or 26 μmol rise within 48 hours. Baseline creatinine will be the median value in the 7 to 365 days before hospitalisation. Absent baseline creatinine was determined based on an eGFR of 75 ml/min/1·72m$^2$ using the CKDepi formula or the admission value whichever was lower.

*Chronic kidney disease*
History of chronic kidney disease (CKD) using baseline eGFR was calculated using last creatinine value available from results earlier than 7 days before hospitalisation. CKD was defined as baseline eGFR below 60 ml/min/1·72m$^2$.

**Table S1:** IMD breakdown by National IMD deciles showing the majority of our study population residing in IMD deciles 1-4.

| Characteristic | n | White, n = 1,805[1] | Black, n = 634[1] | Asian, n = 1,983[1] | Mixed/Other, n = 433[1] | Unknown, n = 678[1] | p-value[2] |
|---|---|---|---|---|---|---|---|
| National IMD Decile | 5,490 | | | | | | <0.001 |
| 1 | | 43 (2.4%) | 29 (4.6%) | 33 (1.7%) | 14 (3.3%) | 21 (3.1%) | |
| 2 | | 368 (21%) | 206 (33%) | 506 (26%) | 118 (28%) | 170 (25%) | |
| 3 | | 468 (26%) | 182 (29%) | 705 (36%) | 125 (29%) | 210 (31%) | |
| 4 | | 261 (15%) | 107 (17%) | 378 (19%) | 74 (17%) | 117 (17%) | |
| 5 | | 154 (8.6%) | 56 (8.9%) | 136 (6.9%) | 41 (9.6%) | 55 (8.2%) | |
| 6 | | 155 (8.7%) | 26 (4.1%) | 84 (4.3%) | 23 (5.4%) | 37 (5.5%) | |
| 7 | | 98 (5.5%) | 8 (1.3%) | 47 (2.4%) | 12 (2.8%) | 21 (3.1%) | |
| 8 | | 104 (5.8%) | 6 (1.0%) | 36 (1.8%) | 11 (2.6%) | 20 (3.0%) | |
| 9 | | 85 (4.7%) | 7 (1.1%) | 32 (1.6%) | 6 (1.4%) | 13 (1.9%) | |
| 10 | | 55 (3.1%) | 2 (0.3%) | 16 (0.8%) | 3 (0.7%) | 6 (0.9%) | |
| Unknown | | 14 | 5 | 10 | 6 | 8 | |

[1]n (%)

[2]Pearson's Chi-squared test

**Table S2:** Rockwood (clinical) and Hospital Frailty Risk (coding-based) scoring within our study cohort.

| Characteristic | n | White, n = 1,805[1] | Black, n = 634[1] | Asian, n = 1,983[1] | Mixed/Other, n = 433[1] | Unknown, n = 678[1] | p-value[2] |
|---|---|---|---|---|---|---|---|
| Rockwood | 2,435 | | | | | | 0.002 |
| 1. Very fit | | 23 (2.2%) | 6 (2.2%) | 20 (2.6%) | 4 (2.5%) | 7 (3.4%) | |
| 2. Well | | 90 (8.7%) | 27 (10%) | 80 (11%) | 29 (18%) | 28 (13%) | |
| 3. Managing well | | 234 (23%) | 77 (29%) | 216 (28%) | 52 (32%) | 49 (24%) | |
| 4. Vulnerable | | 191 (18%) | 56 (21%) | 164 (22%) | 26 (16%) | 43 (21%) | |
| 5. Mildly frail | | 149 (14%) | 33 (12%) | 89 (12%) | 10 (6.1%) | 27 (13%) | |
| 6. Moderately frail | | 199 (19%) | 39 (14%) | 114 (15%) | 26 (16%) | 28 (13%) | |
| 7. Severely frail | | 113 (11%) | 23 (8.6%) | 60 (7.9%) | 11 (6.7%) | 20 (9.6%) | |
| 8. Very severely frail | | 38 (3.7%) | 8 (3.0%) | 13 (1.7%) | 5 (3.1%) | 5 (2.4%) | |
| 9. Terminally ill | | 0 (0%) | 0 (0%) | 2 (0.3%) | 0 (0%) | 1 (0.5%) | |
| Unknown | | 768 | 365 | 1,225 | 270 | 470 | |
| HFRS | 5,518 | | | | | | <0.001 |
| <5 | | 964 (53%) | 377 (60%) | 1,326 (67%) | 315 (73%) | 506 (75%) | |
| 5-15 | | 627 (35%) | 180 (29%) | 509 (26%) | 98 (23%) | 147 (22%) | |
| >15 | | 211 (12%) | 74 (12%) | 144 (7.3%) | 19 (4.4%) | 21 (3.1%) | |
| Score (median) | | 4.3 (1.0, 9.9) | 3.6 (0.5, 8.4) | 2.3 (0.0, 6.5) | 1.8 (0.0, 5.3) | 1.5 (0.0, 4.9) | <0.001 |
| Unknown | | 3 | 3 | 4 | 1 | 4 | |

[1]n (%); Median (IQR)

[2]Pearson's Chi-squared test; Kruskal-Wallis rank sum test

**Table S3:** Age and ethnicity breakdown of First Wave COVID-19 associated hospital admission population residing within Tower Hamlets, Newham or Waltham Forest.

| First Wave n = 1409 | White | Black | Asian | Mixed | Other | Unknown |
|---|---|---|---|---|---|---|
| *Age 16 to 19* | 2 | 0 | 6 | 0 | 0 | 1 |
| *Age 20 to 24* | 3 | 3 | 6 | 1 | 0 | 2 |
| *Age 25 to 29* | 5 | 3 | 24 | 0 | 3 | 3 |
| *Age 30 to 34* | 7 | 6 | 25 | 1 | 2 | 6 |
| *Age 35 to 39* | 12 | 8 | 20 | 0 | 8 | 9 |
| *Age 40 to 44* | 12 | 13 | 32 | 0 | 2 | 7 |
| *Age 45 to 49* | 19 | 15 | 44 | 1 | 11 | 7 |
| *Age 50 to 54* | 31 | 23 | 34 | 1 | 8 | 16 |
| *Age 55 to 59* | 40 | 32 | 35 | 1 | 18 | 17 |
| *Age 60 to 64* | 40 | 24 | 47 | 1 | 7 | 11 |
| *Age 65 to 69* | 27 | 19 | 48 | 1 | 8 | 9 |
| *Age 70 to 74* | 50 | 26 | 22 | 1 | 10 | 16 |
| *Age 75 to 79* | 51 | 28 | 22 | 0 | 5 | 10 |
| *Age 80 to 84* | 65 | 39 | 38 | 1 | 3 | 8 |
| *Age 85 and over* | 109 | 33 | 28 | 2 | 7 | 8 |
| ***Total*** | 473 | 272 | 431 | 11 | 92 | 130 |
| ***% Total*** | 33.6% | 19.3% | 30.6% | 0.8% | 6.5% | 9.2% |

**Table S4:** Age and ethnicity Breakdown of Second Wave COVID-19 associated hospital admission population residing within Tower Hamlets, Newham or Waltham Forest.

| Second Wave n = 3891 | White | Black | Asian | Mixed | Other | Unknown |
|---|---|---|---|---|---|---|
| *Age 16 to 19* | 8 | 6 | 16 | 0 | 1 | 2 |
| *Age 20 to 24* | 17 | 7 | 38 | 5 | 8 | 16 |
| *Age 25 to 29* | 26 | 13 | 81 | 1 | 6 | 21 |
| *Age 30 to 34* | 44 | 21 | 107 | 2 | 13 | 31 |
| *Age 35 to 39* | 44 | 23 | 116 | 2 | 18 | 29 |
| *Age 40 to 44* | 50 | 24 | 130 | 1 | 16 | 39 |
| *Age 45 to 49* | 51 | 34 | 137 | 1 | 25 | 38 |
| *Age 50 to 54* | 67 | 39 | 119 | 4 | 25 | 36 |
| *Age 55 to 59* | 79 | 59 | 142 | 3 | 24 | 34 |
| *Age 60 to 64* | 92 | 48 | 173 | 6 | 34 | 35 |
| *Age 65 to 69* | 85 | 39 | 166 | 1 | 27 | 23 |
| *Age 70 to 74* | 125 | 40 | 106 | 1 | 16 | 21 |
| *Age 75 to 79* | 115 | 47 | 95 | 1 | 11 | 17 |
| *Age 80 to 84* | 105 | 52 | 130 | 3 | 13 | 20 |
| *Age 85 and over* | 178 | 33 | 98 | 1 | 13 | 22 |
| ***Total*** | 1086 | 485 | 1654 | 32 | 250 | 384 |
| ***% Total*** | 27.9% | 12.5% | 42.5% | 0.8% | 6.4% | 9.9% |

**Table S5:** Age and sex breakdown of 105,867 Barts Health Emergency Hospital admissions residing within Tower Hamlets, Newham or Waltham Forest during 2013-18 inclusive.

| Barts Health Pre-COVID n = 105,867 | White | Black | Asian | Mixed | Other | Unknown |
|---|---|---|---|---|---|---|
| *Age 16 to 19* | 1161 | 617 | 1529 | 191 | 212 | 247 |
| *Age 20 to 24* | 2768 | 925 | 2341 | 223 | 514 | 560 |
| *Age 25 to 29* | 3846 | 973 | 3080 | 238 | 713 | 779 |
| *Age 30 to 34* | 3706 | 1039 | 3375 | 195 | 681 | 814 |
| *Age 35 to 39* | 3265 | 961 | 3243 | 151 | 565 | 672 |
| *Age 40 to 44* | 2842 | 978 | 2709 | 126 | 502 | 553 |
| *Age 45 to 49* | 3069 | 1218 | 2196 | 119 | 473 | 526 |
| *Age 50 to 54* | 3119 | 1320 | 1932 | 103 | 450 | 509 |
| *Age 55 to 59* | 3155 | 1172 | 1959 | 87 | 392 | 444 |
| *Age 60 to 64* | 2930 | 698 | 1917 | 61 | 355 | 347 |
| *Age 65 to 69* | 3313 | 589 | 1385 | 44 | 279 | 288 |
| *Age 70 to 74* | 3217 | 620 | 1322 | 39 | 232 | 288 |
| *Age 75 to 79* | 3152 | 826 | 1673 | 54 | 224 | 271 |
| *Age 80 to 84* | 3255 | 741 | 1148 | 32 | 180 | 245 |
| *Age 85 and over* | 4742 | 542 | 751 | 32 | 217 | 321 |
| ***Total*** | 47540 | 13219 | 30560 | 1695 | 5989 | 6864 |
| ***% Total*** | 44.9% | 12.5% | 28.9% | 1.6% | 5.7% | 6.5% |

**Table S6:** Age and sex breakdown by ethnicity in the Tower Hamlets, Newham or Waltham Forest population based on the 2011 UK census.

| Tower Hamlet, Newham & Waltham Forest Census N=664295 | White | Black | Asian | Mixed | Other |
|---|---|---|---|---|---|
| *Age 16 to 19* | 12068 | 7400 | 16300 | 2745 | 1957 |
| *Age 20 to 24* | 32548 | 10103 | 31710 | 4036 | 5611 |
| *Age 25 to 29* | 47264 | 10798 | 37870 | 3791 | 7379 |
| *Age 30 to 34* | 39888 | 9823 | 31114 | 2808 | 5458 |
| *Age 35 to 39* | 28682 | 9080 | 22654 | 2089 | 3631 |
| *Age 40 to 44* | 24936 | 11039 | 16200 | 1757 | 3211 |
| *Age 45 to 49* | 21841 | 10002 | 11313 | 1293 | 2148 |
| *Age 50 to 54* | 19002 | 7026 | 10316 | 946 | 1498 |
| *Age 55 to 59* | 15857 | 4003 | 8400 | 549 | 1122 |
| *Age 60 to 64* | 15047 | 2656 | 5262 | 362 | 852 |
| *Age 65 to 69* | 10637 | 2369 | 3602 | 278 | 477 |
| *Age 70 to 74* | 8521 | 2540 | 3965 | 231 | 423 |
| *Age 75 to 79* | 7560 | 1691 | 2542 | 159 | 274 |
| *Age 80 to 84* | 6336 | 894 | 1319 | 99 | 162 |
| *Age 85 and over* | 6378 | 483 | 615 | 67 | 107 |
| ***Total*** | 296565 | 89907 | 203182 | 21210 | 34310 |
| ***% Total*** | 46.0% | 13.9% | 31.5% | 3.3% | 5.3% |

**Table S7:** *ETHICAL* diversity and absolute numbers of ethnic minority patients in comparison to other large UK reports.

| | **OpenSafely First Wave** | | **OpenSafely Second wave (1/9/20 to 31/12/20)** | | | | **SGSS/CHESS (PHE report)** | | **ISARIC (SSRN)** | | **ONS 2/3-28/7/21** | | **ETHICAL-1** | | **ETHICAL-2** | |
|---|---|---|---|---|---|---|---|---|---|---|---|---|---|---|---|---|
| COVID-related Outcome | Death | | Hospital Admission | | Death | | Hospital Admission | | Hospital Admission | | Death | | Hospital Admission | | Hospital Admission | |
| White | 7119 | 65.5% | 12120 | 64.3% | 4874 | 66.2% | 5149 | 79.6% | 25547 | 83.2% | 41855 | 89.6% | 703 | 35.2% | 1805 | 32.6% |
| Black | 250 | 2.3% | 362 | 1.9% | 75 | 1.0% | 323 | 5.0% | 1094 | 3.6% | 1510 | 3.2% | 340 | 17.0% | 634 | 11.5% |
| South Asian | 608 | 5.6% | 2100 | 11.1% | 532 | 7.2% | 581 | 9.0% | 1388 | 4.5% | 2024 | 4.3% | 538 | 27.0% | 1983 | 35.8% |
| Other | 110 | 1.0% | 354 | 1.9% | 61 | 0.8% | 414 | 6.4% | 2664 | 8.7% | 1307 | 2.8% | 156 | 7.8% | 433 | 7.8% |
| Unknown | 2777 | 25.6% | 3919 | 20.8% | 1824 | 24.8% | - | - | - | - | - | - | 259 | 13.0% | 678 | 12.3% |
| Total | 10864 | 100% | 18855 | 100% | 7366 | 100% | 6467 | 100% | 30693 | 100% | 46696 | 100% | 1996 | 100% | 5533 | 100% |

**Table S8:** Age, sex, and mortality by detailed ethnicity breakdown.

| | African, n = 262[1] | Any other Asian background, n = 379[1] | Any other Black background, n = 133[1] | Any other ethnic group, n = 358[1] | Any other mixed background, n = 22[1] | Any other White background, n = 507[1] | Bangladeshi, n = 926[1] | British, n = 1,269[1] | Caribbean, n = 239[1] | Chinese, n = 30[1] | Indian, n = 301[1] | Irish, n = 29[1] | Not known, n = 509[1] | Not stated, n = 169[1] | Pakistani, n = 377[1] | White and Asian, n = 4[1] | White and Black African, n = 13[1] | White and Black Caribbean, n = 6[1] |
|---|---|---|---|---|---|---|---|---|---|---|---|---|---|---|---|---|---|---|
| Age[1] | 55 (43, 65) | 56 (43, 70) | 58 (44, 68) | 58 (45, 70) | 53 (36, 66) | 52 (39, 68) | 54 (40, 68) | 74 (61, 83) | 72 (58, 81) | 62 (44, 73) | 66 (52, 77) | 74 (62, 83) | 52 (39, 67) | 63 (49, 74) | 60 (42, 71) | 51 (48, 56) | 57 (33, 61) | 71 (53, 83) |
| Male | 130 (50%) | 225 (59%) | 66 (50%) | 206 (58%) | 10 (45%) | 273 (54%) | 484 (52%) | 655 (52%) | 109 (46%) | 11 (37%) | 165 (55%) | 18 (62%) | 284 (56%) | 91 (54%) | 206 (55%) | 3 (75%) | 5 (38%) | 3 (50%) |
| 30-day Mortality | 27 (10%) | 54 (14%) | 19 (14%) | 44 (12%) | 2 (9.1%) | 59 (12%) | 166 (18%) | 305 (24%) | 57 (24%) | 4 (13%) | 76 (25%) | 6 (21%) | 62 (12%) | 30 (18%) | 59 (16%) | 1 (25%) | 0 (0%) | 3 (50%) |

[1]Median (IQR); n (%)

[2]Kruskal-Wallis rank sum test

**Fig S1:** Patients considered in second wave *ETHICAL* study 1/9/20 to 17/2/21. Criteria: aged ≥16 first emergency admission to one of the four Barts Health acute hospitals with a positive hospital COVID-19 PCR test during or in the two weeks prior to admission. NUH: Newham University Hospital.

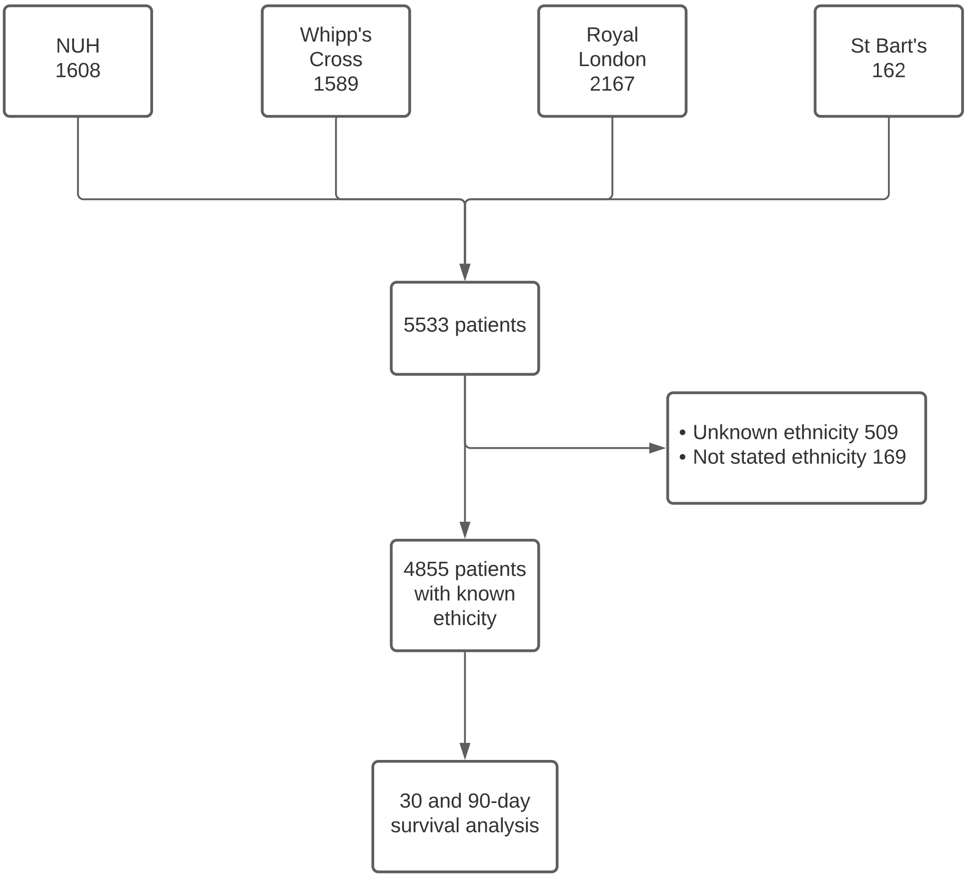

**Fig S2:** Daily COVID-19 associated emergency admission during our study period.

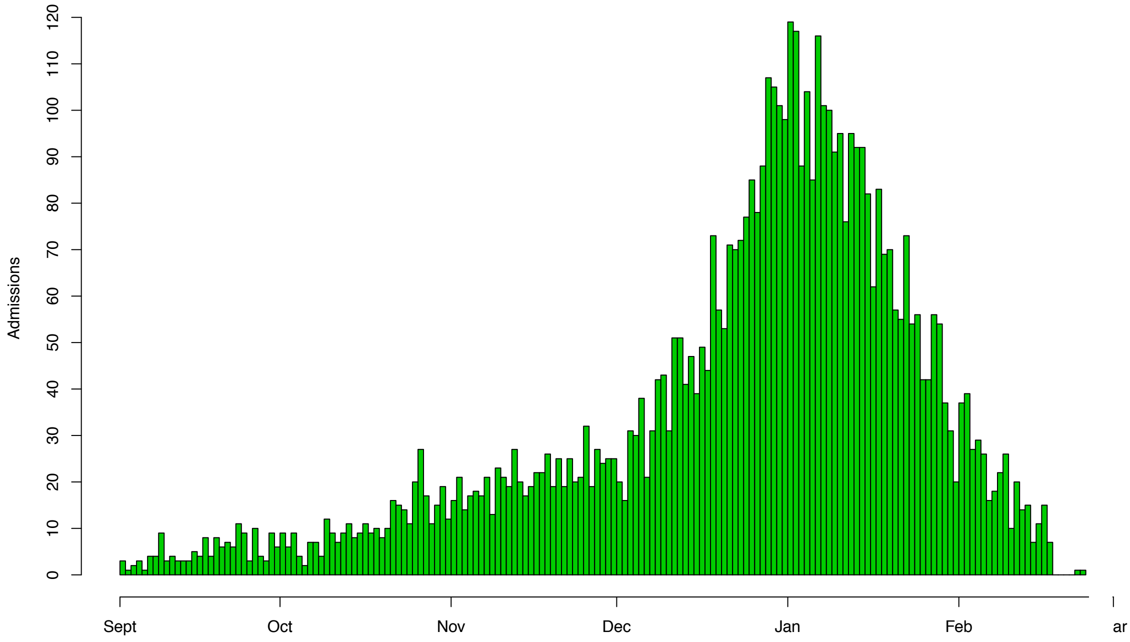

**Fig S3:** Cox-proportional Hazard analysis included covariates: Age, Sex, Charlson Index, Smoking history, Obesity, IMD (quintiles of local study population) for survival over 30 days in the second wave *ETHICAL* cohort.

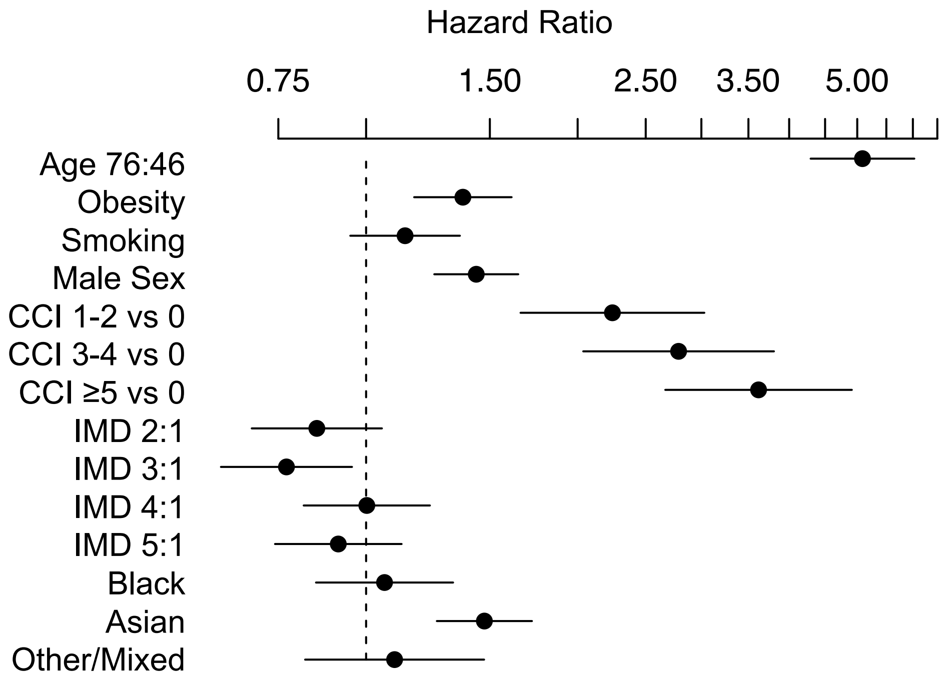

**Fig S4:** Cox-proportional Hazard analysis included covariates: Age, Sex, Hospital Frailty Risk Score, Smoking history, Obesity, IMD (quintiles of local study population) for survival over 30 days in the second wave *ETHICAL* cohort.

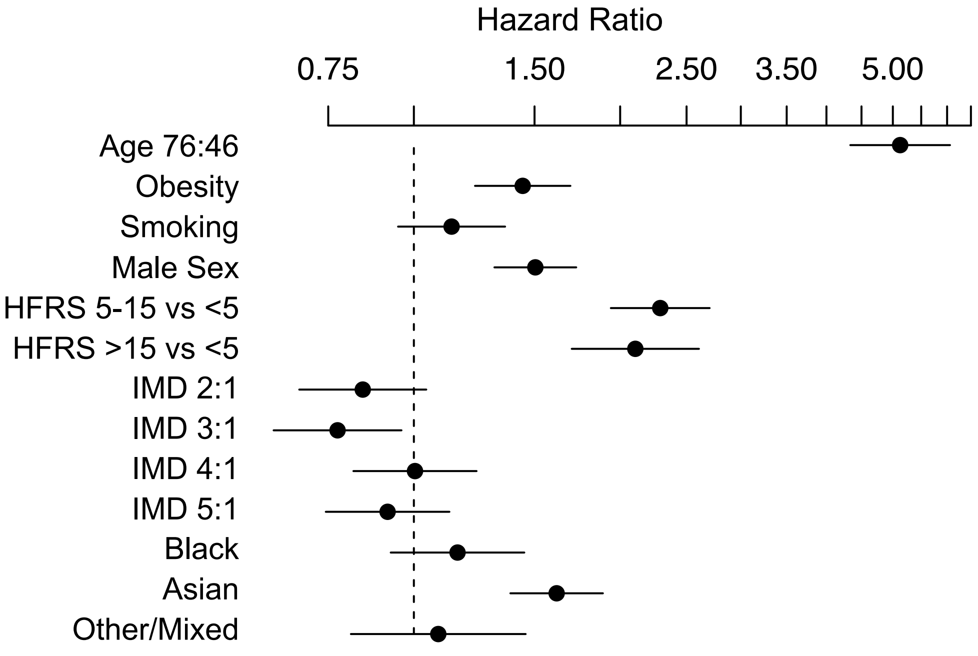

**Fig S5:** Logistic regression model for risk of ICU admission during first COVID associated hospitalisation in second wave adjusted for COVID-19 risk factor comorbidities.

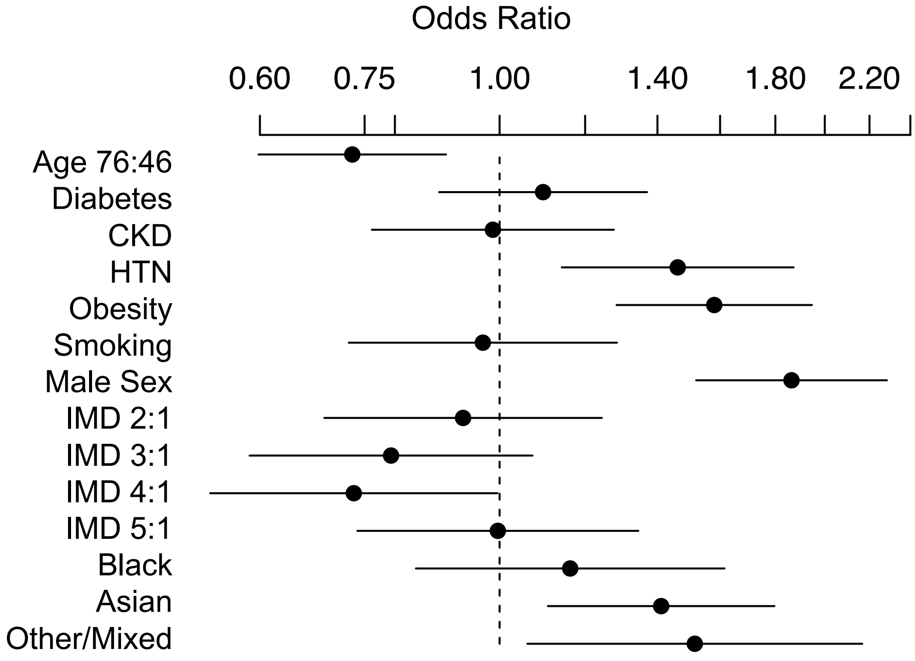

**Fig S6:** Cox-Hazard analysis (unstratified) for survival of first wave patients to 12 months.

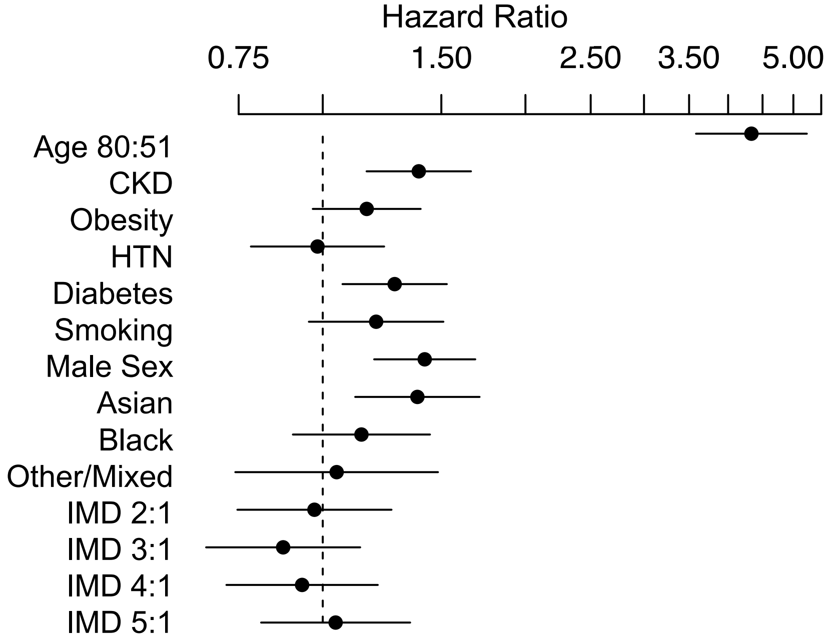
